# A statistical analysis of pulse transit time captured using pressure sensors at the human radial artery of the wrist

**DOI:** 10.64898/2026.05.14.26353264

**Authors:** Sukesh Rao M, Dariush Khezrimotlagh

**Author notes:** **Corresponding author:** Sukesh Rao M, Department of Electronics and Communication Engineering, NMAM Institute of Technology (NMAMIT), Nitte (Deemed to be University), Nitte, Karnataka, India.

## Abstract

Non-invasive wrist pulse monitoring has been integrated into various medical systems for cardiovascular assessment. However, different definitions of pulse transit time are used in the literature, and their statistical behavior when measured locally at the wrist using pressure sensors has not been systematically examined. Wearable wristbands designed to measure pulse transit time (PTT) have emerged as valuable tools for evaluating cardiac activity. While several algorithms have been developed to predict blood pressure using PTT, it is well recognized that PTT and its inverse parameter, pulse wave velocity (PWV), exhibit temporal variability. In this study, PTT was explicitly measured at the wrist’s radial artery to investigate its statistical variation and relationship with different arterial pressures. The experiment exhibits two distinct methodologies for PTT computation using onset-based and peak based measurements. Data were recorded across five cuff pressure levels at 20, 40, 60, 80, and 100 mmHg using the pulse pressure sensor (PPS). PTTonset time shows lower coefficient of variation as compared to PTTpeak time within the 100 mmHg pressure range. The weak correlation coefficient is recorded between PTT values. However, dynamic time warping (DTW) analysis revealed a notable similarity in the time series of PTTonset and PTTpeak, regardless of the applied pressure level. For the multi participant dataset, the mean DTW distances ranged from 0.029 to 0.046 across the tested cuff pressures, illustrating consistent similarity between PTTonset and PTTpeak over time. The objective of this study is to examine the statistical behavior, stability, and temporal similarity of the two commonly used PTT definitions when measured at the radial artery using pressure sensors. Statistical analysis shows consistent differences between the two PTT definitions across participants. PTTonset shows lower variation than PTTpeak. However, PTTpeak requires simpler computation and produces fewer detection errors, while PTTonset provides lower statistical variation.

## 1. Introduction

Wrist pulse monitoring is a fundamental practice across various medical systems for disease diagnosis. Each pulse beat measured at an artery carries valuable information regarding the cardiovascular health. Pulse wave characteristics can be assessed at multiple anatomical locations, including the carotid artery (neck) [1], brachial artery (inner elbow), radial artery (wrist) [2-3], femoral artery (groin), popliteal artery (behind the knee), dorsalis pedis artery (top of the foot), and posterior tibial artery (below the ankle bone). Extensive research has been conducted to acquire pulse signals from different arterial sites, with the wrist emerging as a prominent location for wearable health monitoring devices [4-9].

Key physiological parameters, such as pulse rate, blood pressure (BP), and blood oxygen saturation (SpO2), are predominantly estimated from the radial artery using wearable devices. However, ensuring the accuracy of these measurements is a significant challenge. Non-invasive sensors, including photoplethysmography (PPG) and pressure sensors, are commonly employed for BP estimation via the radial artery [10-11]. Two principal methodologies are utilized for this purpose: (i) volumetric changes in blood flow at the radial artery [12-16] and (ii) estimation based on pulse arrival time (PAT) or pulse wave velocity (PWV) [16-26].

PAT is defined as the time delay between the Q-point of the electrocardiogram (ECG) and the peak of the corresponding pulse measured at a specific arterial site [11], [27]. In contrast, the pulse transit time (PTT) differs from PAT in terms of the measurement approach and underlying signal dynamics. PTT refers to the time delay of a pulse wave propagating between two arterial sites along the radial artery [11]. It can be measured efficiently using PPG or pressure sensors. Notably, signal quality and extracted features vary between PPG-based and pressure-based pulse acquisition systems [28]. Pressure sensors often provide superior signal fidelity, enabling enhanced feature extraction capabilities.

According to the work of Thomas Young et al. the PTT is likely to depend on arterial stiffness, density of the blood and wall thickness [29]. The theoretical basis for the blood flow in the artery was modelled, and it was formally known as Moens-Korteweg equation (Equation 1). According to equation the pulsatile flow of the blood is because of artery wall thickness, elastic modulus of the vessel and internal pressure.

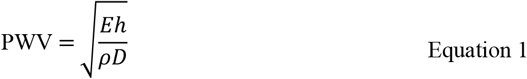

where *E* is the Young’s modulus of the arterial wall, *h* is the wall thickness, *D* is the arterial diameter and *ρ*is the density of blood. Since PTT is inversely related to pulse wave velocity (PWV) as stated in the Equation 2.

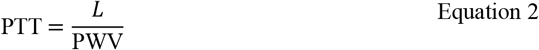

where *L* is denotes as length of the artery.

The young’s formulation implies that the elasticity of the mainly dependent on the pressure gradient leading to a nonlinear relationship between blood pressure and PTT [30]. This indicates that as the blood pressure increases, PWV increases and arterial walls become stiffer. Previous studies have used both peak and onset points of the pulse waveform to compute pulse transit time. While both definitions are widely used, limited attention has been given to how these two measurements behave statistically when measured locally along the radial artery using pressure sensors. In particular, the relative stability of the two definitions, their detection reliability, and their temporal similarity under controlled pressure conditions have not been systematically examined.

In this study, a sensor setup was developed to capture pulse signals at different applied pressure levels to investigate PTT variations. A statistical analysis was performed to examine PTT variation measured using pressure sensor at different cuff pressure. PTT is computed using pulse peak and pulse onset-based approach. PTT obtained using these two different approaches are examined with statistical methods. The insights obtained from this analysis are critical for developing mathematical models to predict blood pressure and characterize blood flow patterns in arteries. The study focuses on comparing the statistical behavior of peak and onset definitions of PTT, including their variability, detection reliability, and temporal similarity across different cuff pressures and across multiple participants.

The rest of this study is organized as follows. Section 2 illustrates the methodology and the measurement of PTT.

## 2. Methodology

Pulse measurements were obtained using an inflatable wristband worn on the left or right hand. The wristband was equipped with a cuff pressure sensor (CPS) for monitoring the internal cuff pressure, along with two pulse pressure sensors (PPS1 and PPS2) positioned to capture pulse waveforms. The sensor configuration and placement are illustrated in Figure 1.

**Figure 1:**
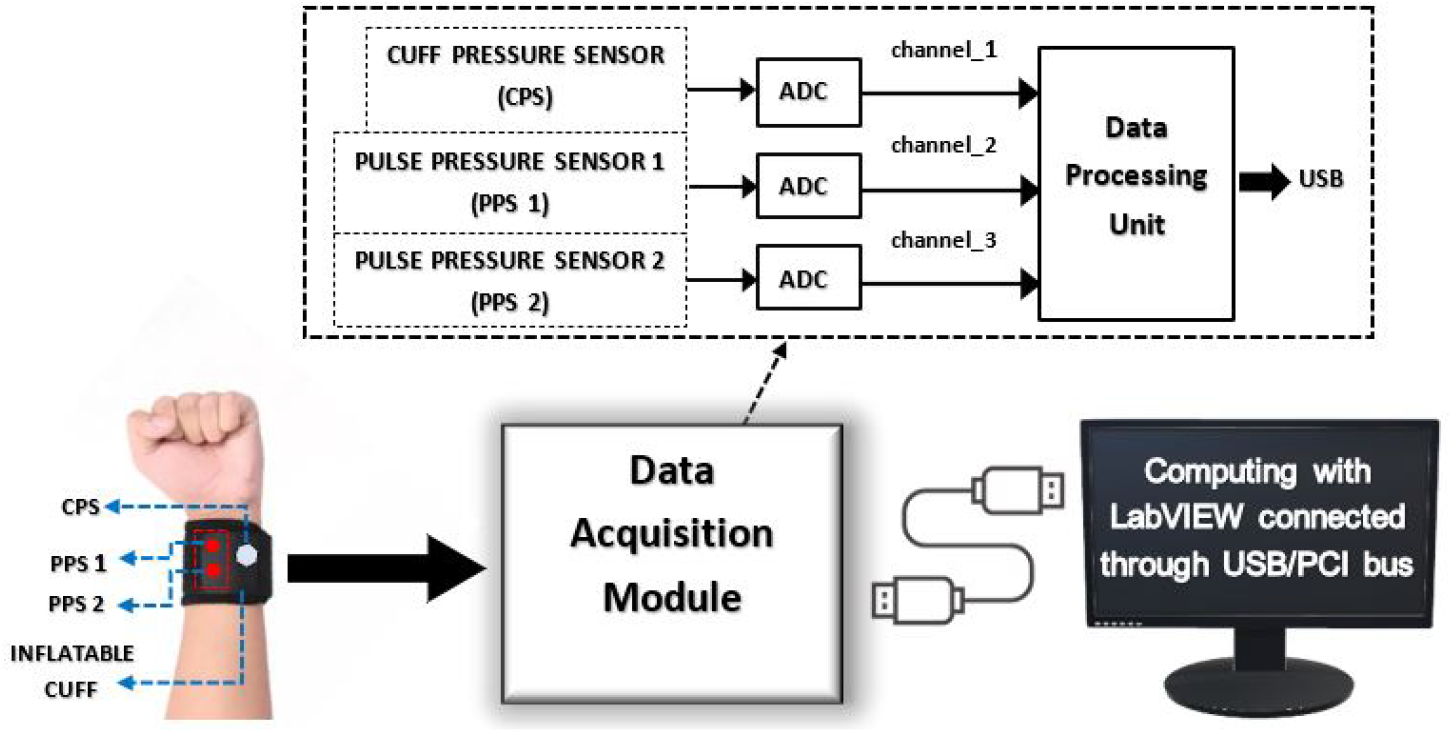
Block diagram of pulse acquisition and computation system.

Recordings were performed at five different cuff pressure levels mainly at 20 mmHg, 40 mmHg, 60 mmHg, 80 mmHg, and 100 mmHg. At each pressure level, pulse waveforms were simultaneously captured from both PPS1 and PPS2 while the subject remained in seated position.

The sensors are connected to acquire three analog signals, one from the cuff pressure sensor and two from pulse pressure sensor. These signals were digitized using USB-6003 data acquisition (DAQ) module via analog to digital converter (ADC) channels. Data acquisition and signal processing were implemented using a LabVIEW virtual instrument (VI). The LabVIEW tool is used to record the pulse waveform data from the participants and stored in a spreadsheet format for further analysis.

The sensor setup used in this study consists of two PPS positioned 2.5 cm apart and mounted on the radial artery of the wrist, as shown in Figure 1. This short separation distance is intended to capture local pulse waveform propagation along the radial artery rather than estimate global arterial pulse wave velocity. The 2.5 cm distance is the optimum distance that can fit two sensors when mounted with two fingers of the physician [11]. This spacing also provides sufficient propagation distance to measure the delay of the pulse waveform while maintaining practical placement on the wrist.

### 2.1. Measurement of PTT

Radial artery of the wrist provides the easy access to measure the pulsatile flow of the blood. Pressure sensors are placed at the radial artery of the wrist to capture the pulsatile flow of the blood. A typical pulsatile flow of the blood is shown in Figure 2, representing the systolic and diastolic phase along with other features [11].

**Figure 2:**
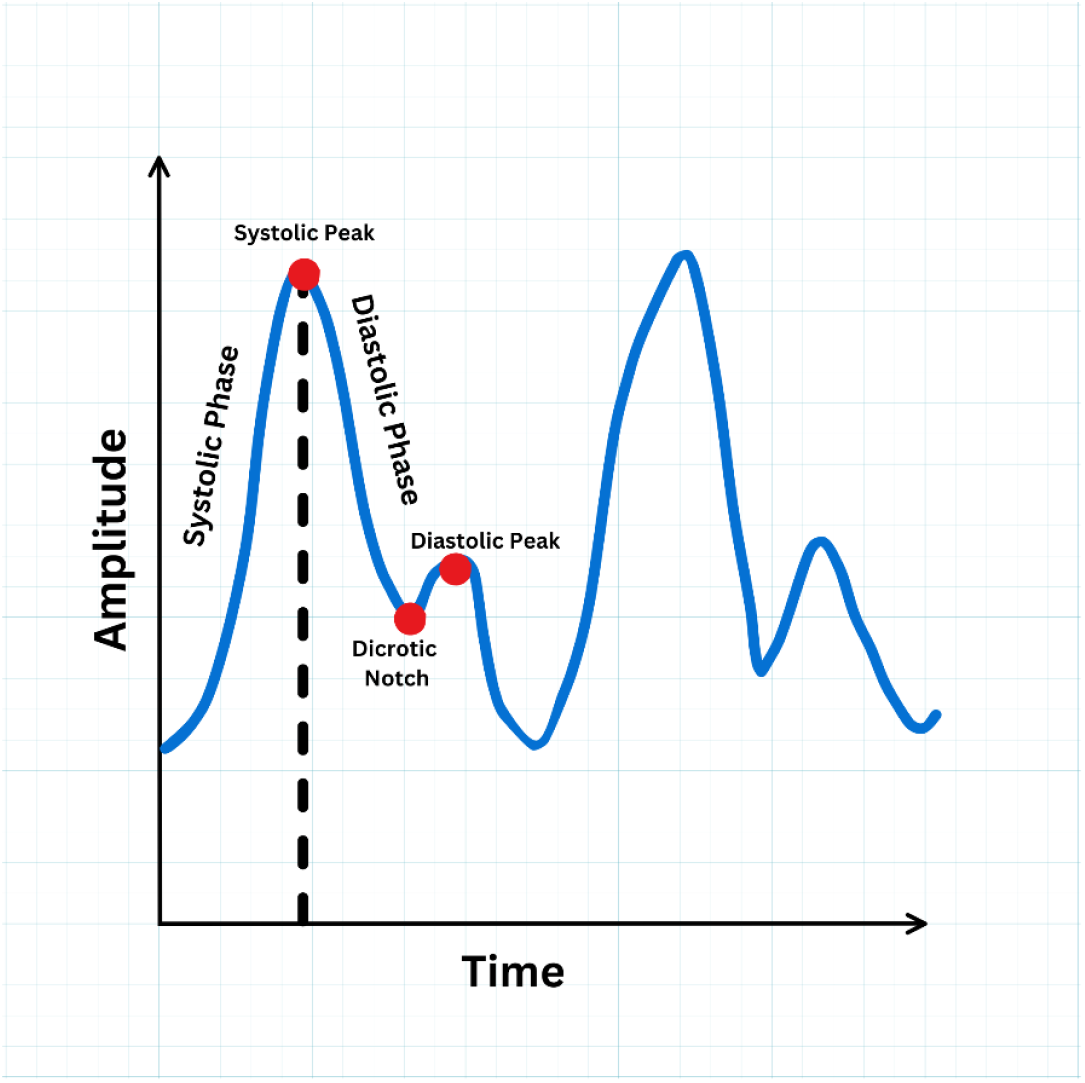
Representation of basic pulse wave features.

A measurement made using two sensors at two different given points in the radial artery can provide the significant information such as pulse wave velocity and pulse transit time. PTT provides the transition time of the blood flow from one point to the other in the cardiac cycle. Two pressure sensors (Sensor 1 and Sensor 2) are placed at the radial artery of wrist as shown in Figure 3(a) then the pulse acquired from the sensors are pulse 1 and pulse 2 respectively. The time domain signals of the pulse 1 and pulse 2 are shown in Figure 3(b) with marking of the PTT information. PTT can be computed based on the time index of peak amplitude of the pulse or onset time of the pulse. In the experimental setup the pulse wave reaches the upstream sensor first and then the downstream sensor along the radial artery. In this study the signal from sensor 2 precedes the signal from sensor 1, which is reflected in the timing diagram shown in Figure 3(b). The PTT from the peak time index is computed as difference between the peak time index of pulse 1 and peak time index of pulse 2. In other case, PTT is computed using onset time method [31]. Onset time is the starting point of the pulse. Computing the onset time of a pulse is described in detail in the subsection 2.1.1. **Error! Reference source not found**. and **Error! Reference source not found**. can be used to compute the PTT using peak time index and onset-based method respectively.

**Figure 3:**
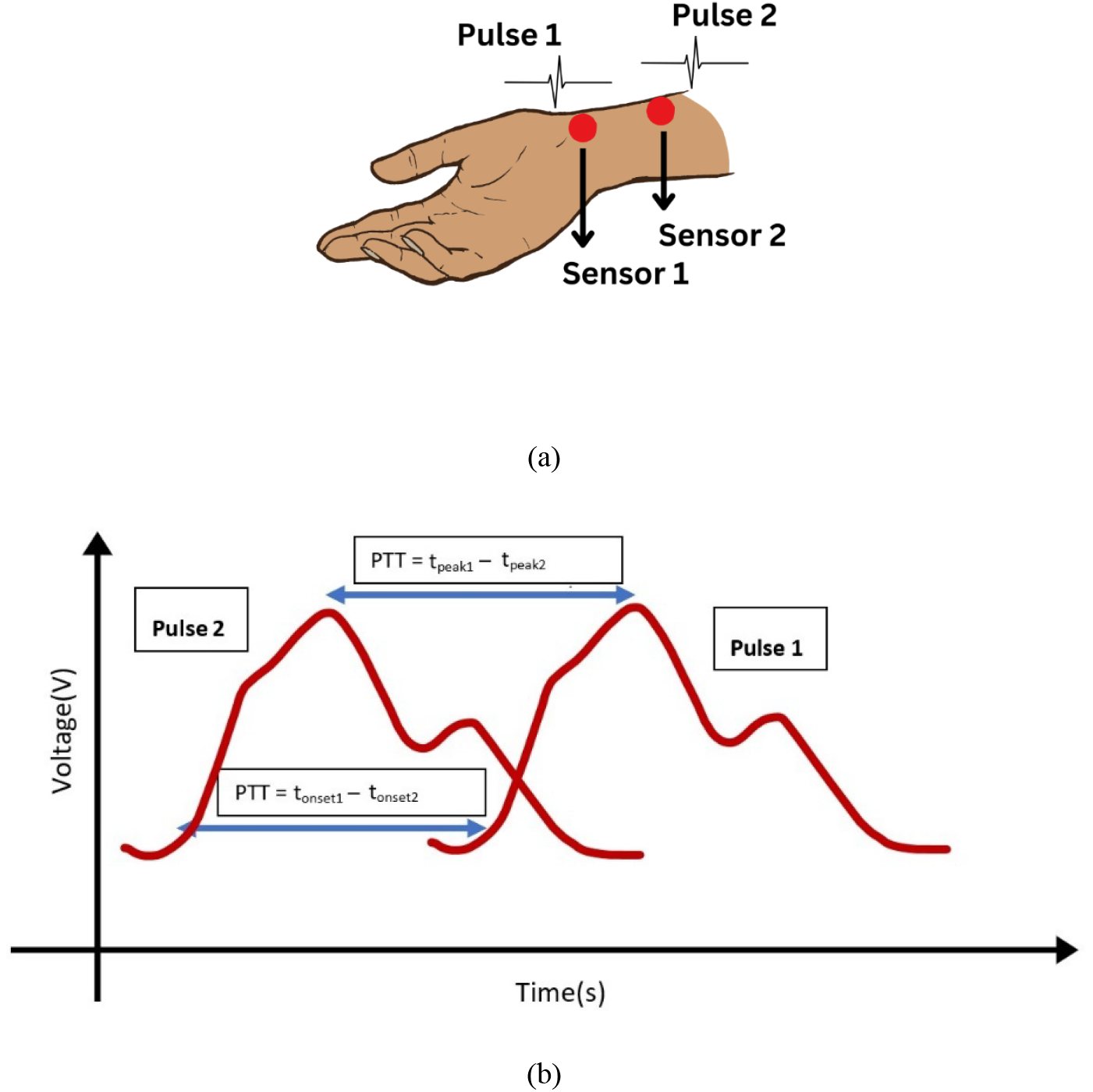
a) Placement of sensor 1 and sensor 2 at radial artery; b) Basic computation of PTT in a cardiac cycle from Sensor 1 and Sensor 2.

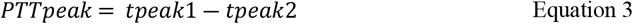

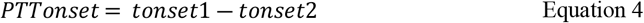

#### 2.1.1. Method to compute onset time of a pulse

It is hard to locate the starting point of the pulse in a cardiac cycle. In practice, onset detection is more sensitive to waveform distortion and motion artifacts than peak detection, which motivates the comparison between onset and peak definitions in this study. There could be more ambiguity in defining the starting point of a pulse due to motion artifact or inconsistent movement of the pulse duration. Hence, an indirect method is used by constructing tangential lines at two different significant markers of the pulse cycle. At first, a tangential line is drawn with the minimum point of the pulse. In the next process, 1st order derivative is computed for the pulse samples [31]. The maximum value of the 1st order derivative is marked on the pulse signal. The point where tangential line of minimum and 1st order maximum derivative intercepts is considered as an onset point of the pulse. Figure 4 represents the actual marking of the tangential lines on the pulse. The procedure is repeated for every pulse cycle of pulse 1 and pulse 2 for predicting the pulse onset time. The difference between the onset time of pulse 1 and pulse 2 is treated as PTT Onset.

**Figure 4:**
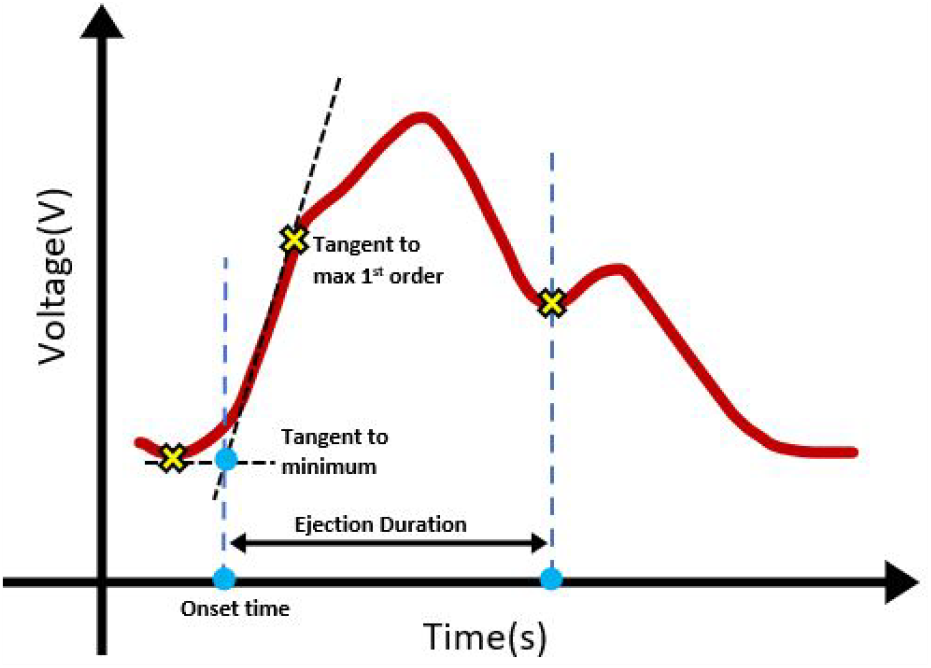
Locating onset time on the pulse.

### 2.2. Calibration of CPS

Pulse waveform data from are recorded at different cuff pressure of the inflatable wristband as in Figure 1. It is essential to measure the absolute pressure of the wrist band to mark the pulse waveform data. Calibration is made using a standard sphygmomanometer setup as shown in Figure 5. Curve fitting method is used to map the analogue voltage (Vout) of the CPS to mmHg scale. The gain value of the instrumentation amplifier is kept at 22.02 because of standard passive components used in the design. The gain value is suitable to get the signal strength within the required Input voltage range for the ADC.

**Figure 5:**
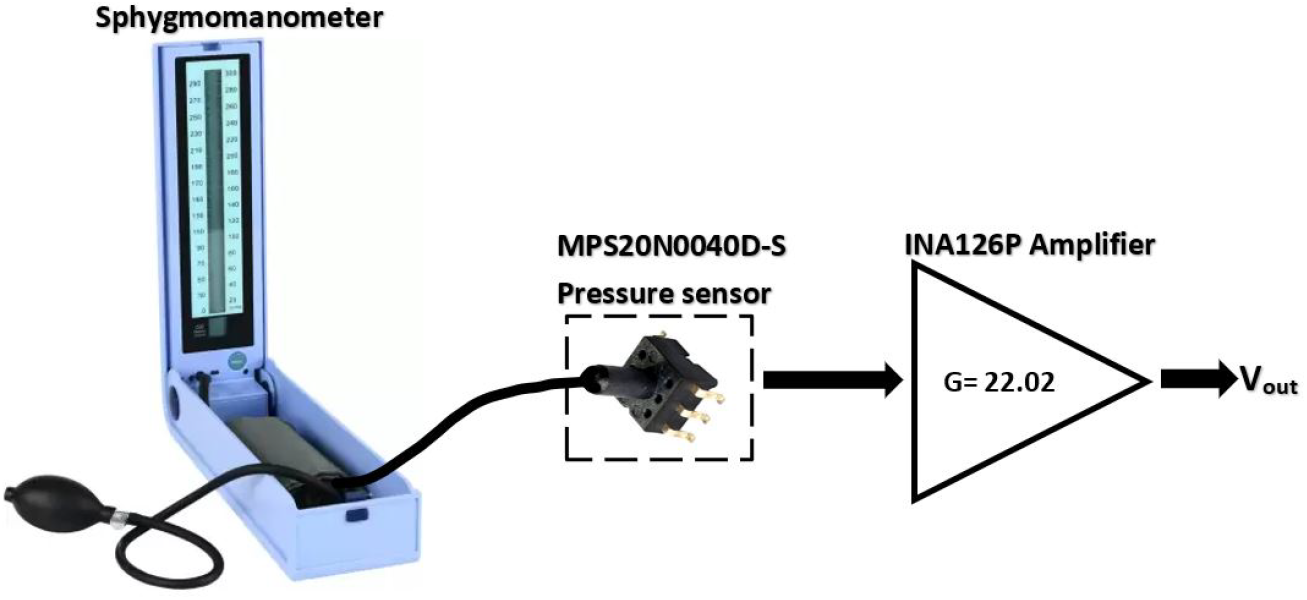
Calibration process if cuff pressure sensor placed inside wrist band.

### 2.3. Construction of sensor module to measure PTT

Pressure sensors are fabricated on to a customized PCB for the easy measurement of the pulsatile flow at radial artery of wrist. Figure 6 provides the detailed schematic and developed sensor modules for the measurement of the pulse for the experimental purpose.

**Figure 6:**
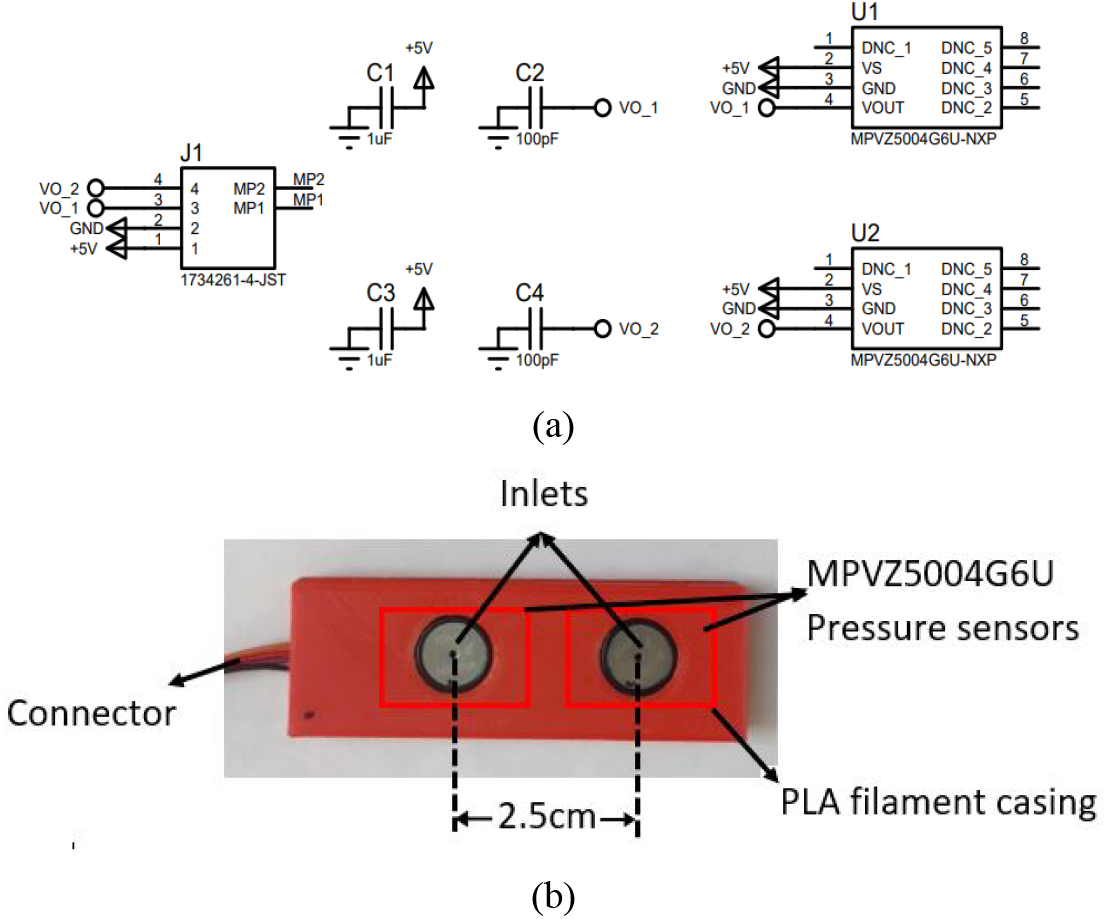
(a) Schematic Design for MPVZ004G6U; (b) Complete PCB with encloser for the MPVZ5004G6U sensors.

### 2.4. Data collection process and analysis

The study was performed in two phases: an exploratory phase using a single participant for feature extraction and algorithm setup, and a validation phase involving 18 participants for statistical analysis. In the exploratory phase, initial experiment is performed with single participant and data recoding is collected for 1 minute duration using the sensor setup. The data from single participant is used for initial investigation and features extraction. Pulse recording from the single participant is used to obtain the PTTonset and PTTpeak time values. In the validation phase, statistical analysis is made on the diverse set of the reading from the 18 participants to validate the study.

The pulse acquisition process is carried out by planning the pressure sensor in each participant as explained in the section 2.2. Further recordings are obtained from 18 healthy participants in the relaxed and sitting position for about 15 seconds each. Data recording is performed using DAQ module and stored in the spreadsheet for the analysis in the MATLAB simulation tool.

An algorithm is developed to capture the PTTonset and PTTpeak from each dataset stored in the spreadsheet and the same algorithm was applied to all 18 participants. The mean values of PTTpeak and PTTonset are tabulated for observing the variation at different pressure level. Also, valid positive and negative count rate of PTTonset and PTTpeak are separately tabulated. In addition, the coefficient of variation is used to evaluate measurement stability across pressure levels, correlation is used to examine linear association between the two PTT definitions, and discrete time wrapping (DTW) technique is used to evaluate similarity of their temporal patterns. The results are further explored with 18 participants for the statistical analysis. The Jarque-Bera (JB) Test [32] is a statistical test used to check whether a given sample of data follows a normal distribution. In this study the test is used to determine whether parametric or nonparametric statistical comparisons should be used in the subsequent analysis.

## 3. Implementation and Results

It was evident from the past experimental result that pulse activity at wrist is found to be predominant and easy to measure [32-33]. In this study, a new attempt is made to measure the PTT at different pressure levels at the radial artery of wrist. The results are presented in two phases: an exploratory phase using a single participant and a validation phase using 18 participants. In other words, the behavior of PTTpeak and PTTonset is first examined for the initial subject in section 3.1, and the observations are later validated using the dataset from 18 participants in section 3.2. The purpose of the exploratory phase is to illustrate the behavior of the measurement system and verify the signal processing procedure before examining the multi participant dataset.

### 3.1. The exploratory phase

Initial experiment is conducted to check the PTT information at different cuff pressure varying from 20 mmHg to 100 mmHg levels for a single participant. A digital filtering technique is used to filter the high frequency noises with a cutoff frequency 10 Hz. The signals were filtered using a low pass finite impulse response filter with sampling frequency 1000 Hz and filter order 500. The signals obtained from the sensors are stored in the excel sheet for the pulse entries corresponding to 20 mmHg, 40 mmHg, 60 mmHg, 80 mmHg, and 100 mmHg pressure levels labelled as sheet 20, sheet 40 and so on. Figure 7 represents the result of the signal after filtering process for the initial subject. Each sheet will contain the recording of the pulse waveform data for about one minute at a sampling frequency 1000 Hz. Hence there are about 60,000 samples in each sheet. It is more evident that the pulse at 20 mmHg to 60 mmHg are more fluctuating type, whereas pulses are steadier and more stable beyond 60 mmHg level. It is also observed that the pulses are more predominating at 60 to 100 mmHg, and the pulse signals appear more stable at higher cuff pressures. The signal after filtering is used to predict the PTT and other pulse features for the further analysis. As it is explained in subsection 3.2, these steps are also done for the rest of 18 participants.

**Figure 7:**
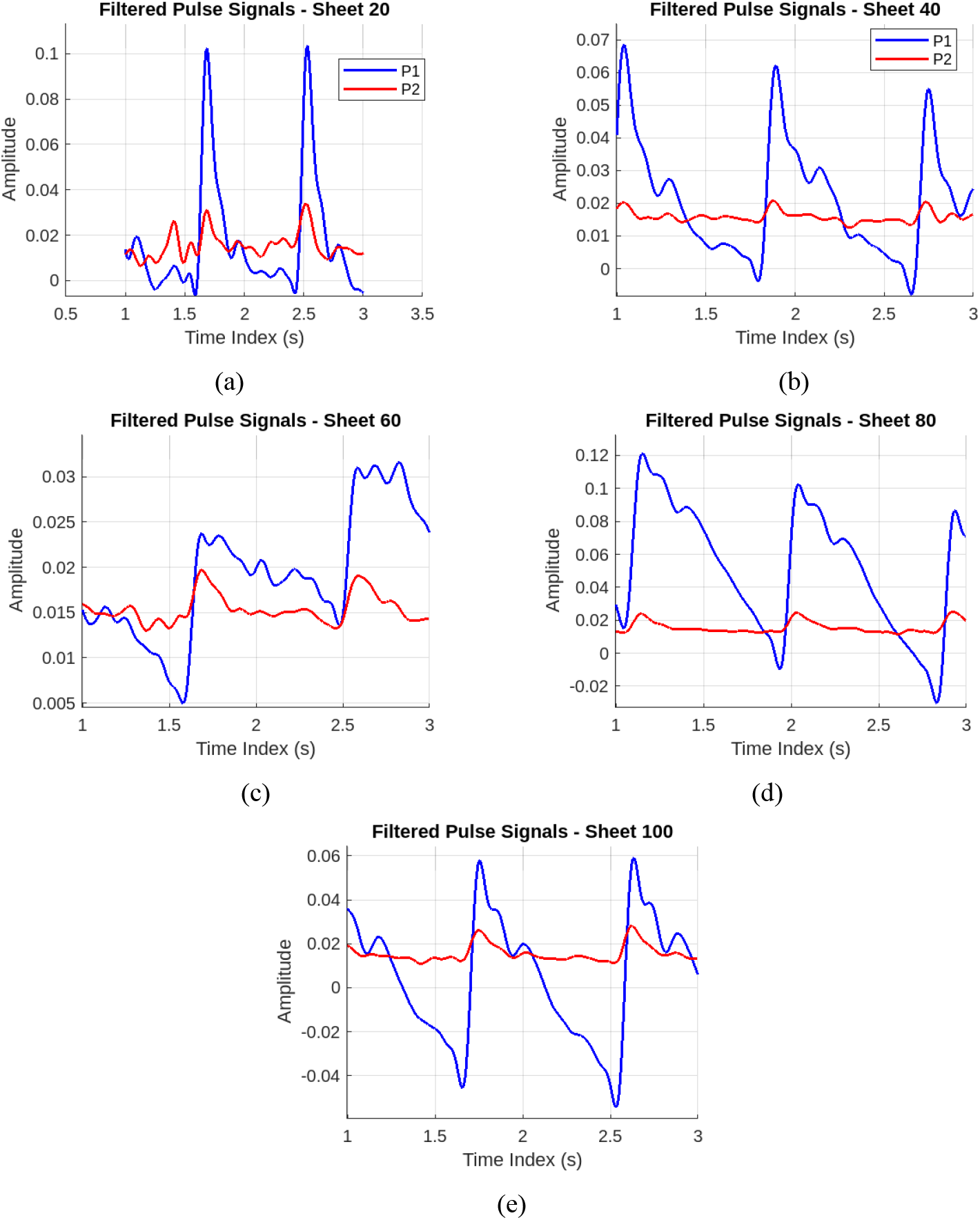
Pulse signal after the preprocessing process for 20 mmHg sheet to 100 mmHg sheet.

#### 3.1.1. PTT variation in peak and onset method

The pulse corresponded to sheet 20 to sheet 100 from the initial subject are segmented at a fixed duration for the easy computation of the PTT. The algorithm for computing the PTTpeak and PTTonset are developed in the MTALAB tool. Local maxima were detected by checking whether each sample is the absolute maximum within a window of 100 samples before and 100 samples after the candidate point. According to Figure 4, P1 peak or onset should appear after P2. However, due to motion artifact and turbulent nature of the blood flow the sequence may change. To observe the sequence for valid computation of PTT, a count information is recoded to find the true peak (P2 first and then P1) and is named as positive count. A negative count indicates the false peak (P1 first and then P2) and is named as negative count. In the implementation, valid PTT pairs were formed when a detection from sensor 2 was followed by the corresponding detection from sensor 1 in consecutive rows, and PTT was computed as the time difference between the two detections. Similar computation of positive and negative count is made for onset of P1 and P2 as well. The actual PTT is computed using the true peak having positive count only and negative count onset/peak must be discarded. Table 1 provides the positive peak and negative peak count for the pulse sheet 20 to sheet 100 for the initial subject. Similarly, Table 2 provides the count information for onset values.

**Table 1:** Peak values count of the pulse waveform.

| *Table 1: Peak values count of the pulse waveform.* |  |  |  |
| --- | --- | --- | --- |
| Pressure (mmHg) | Negative Count | Positive Count | Total Count |
| 20 | 0 | 69 | 69 |
| 40 | 0 | 72 | 72 |
| 60 | 0 | 47 | 47 |
| 80 | 0 | 52 | 52 |
| 100 | 0 | 46 | 46 |

**Table 2:** Onset values count for the pulse waveform.

| *Table 2: Onset values count for the pulse waveform.* |  |  |  |
| --- | --- | --- | --- |
| Pressure (mmHg) | Negative Count | Positive Count | Total Count |
| 20 | 24 | 45 | 69 |
| 40 | 9 | 63 | 72 |
| 60 | 7 | 40 | 47 |
| 80 | 0 | 52 | 52 |
| 100 | 0 | 46 | 46 |

**Table 3:** Mean, standard deviation and coefficient of variation of PTTpeak and PTTonset.

|  |  |  |  |  |
| --- | --- | --- | --- | --- |
| Pressure (mmHg) | PTT<sub>peak</sub> (Mean ± SD) | PTT<sub>onset</sub> (Mean ± SD) | PTT<sub>peak</sub> CoV | PTT<sub>onset</sub> CoV |
| 20 | 0.0101 ± 0.0038 | 0.0066 ± 0.0024 | 37.62% | 35.82% |
| 40 | 0.0130 ± 0.0040 | 0.0069 ± 0.0016 | 31.24% | 22.66% |
| 60 | 0.0116 ± 0.0080 | 0.0062 ± 0.0013 | 68.93% | 20.96% |
| 80 | 0.0150 ± 0.0060 | 0.0079 ± 0.0013 | 39.84% | 15.80% |
| 100 | 0.0115 ± 0.0046 | 0.0080 ± 0.0018 | 40.09% | 22.36% |

In Table 1 and Table 2, the maximum pulse count recorded is 72 for the 60 second recording consisting of 60,000 samples at a sampling rate of 1 kHz. The count value reflects the number of detected pulse cycles in the analyzed dataset and is used only for verifying the valid peak and onset detections. Since the recording duration is 60 seconds, the total count of 72 also corresponds approximately to the heart rate of the participant during that recording. If values less than 72 indicate, certain pulse segments are discarded in the programming due to threshold level in selecting the valid pulse waveform. The valid pulse total counts exactly match in Table 1 and Table 2, indicates only those pulses are considered for verification of negative and positive count. It is found that peak based PTT produces fewer detection errors in the pulse waveform with no entries in the negative count. Negative counts occur only in the onset detection method due to ambiguity in identifying the pulse onset. For example, at 20 mmHg the onset method produced 24 negative detections out of 69 pulses, whereas the peak method produced no negative counts. The values may vary in different iteration; however, PTTonset computation may lead to negative pulse count due to motion artifact. To reduce false detections, peaks with amplitudes below the average peak amplitudes of the recorded signals were removed before pairing pulses between the two sensors.

The PTTpeak and PTTonset is computed using **Error! Reference source not found**. and **Error! Reference source not found**. respectively for the listed positive count obtained in the Table 1 and Table 2. Figure 8(a) and 8(b) show the overall smoothed and trimmed variation of PTTpeak and PTTonset. A 5-point moving average is performed to observe the variation in the PTT.

**Figure 8:**
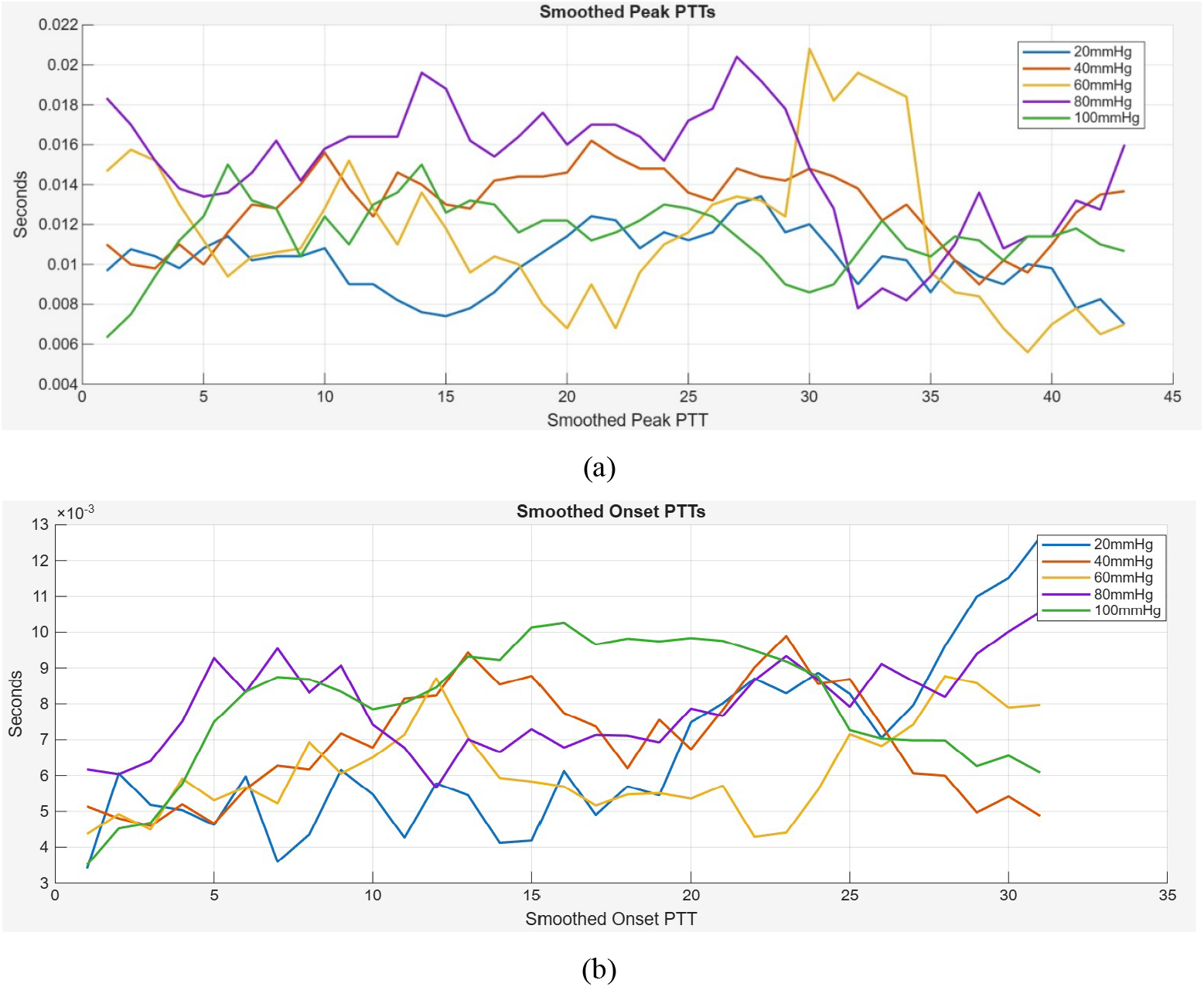
(a) PTTpeak values with 5 point moving average; (b) PTTonset values with 5 point moving average values.

The positive values of PTTpeak and PTTonset are considered to compute the mean, standard deviation and coefficient of variation as listed in Table 3. From Table 3, it can be observed that the PTTonset less variation in contrast to PTTpeak. Consistently lower variation is observed for PTTonset values across pressure levels in this dataset. The normalized peak and onset PTT variation of corresponding to different pressure levels are shown in Figure 9 for the initial subject.

**Figure 9:**
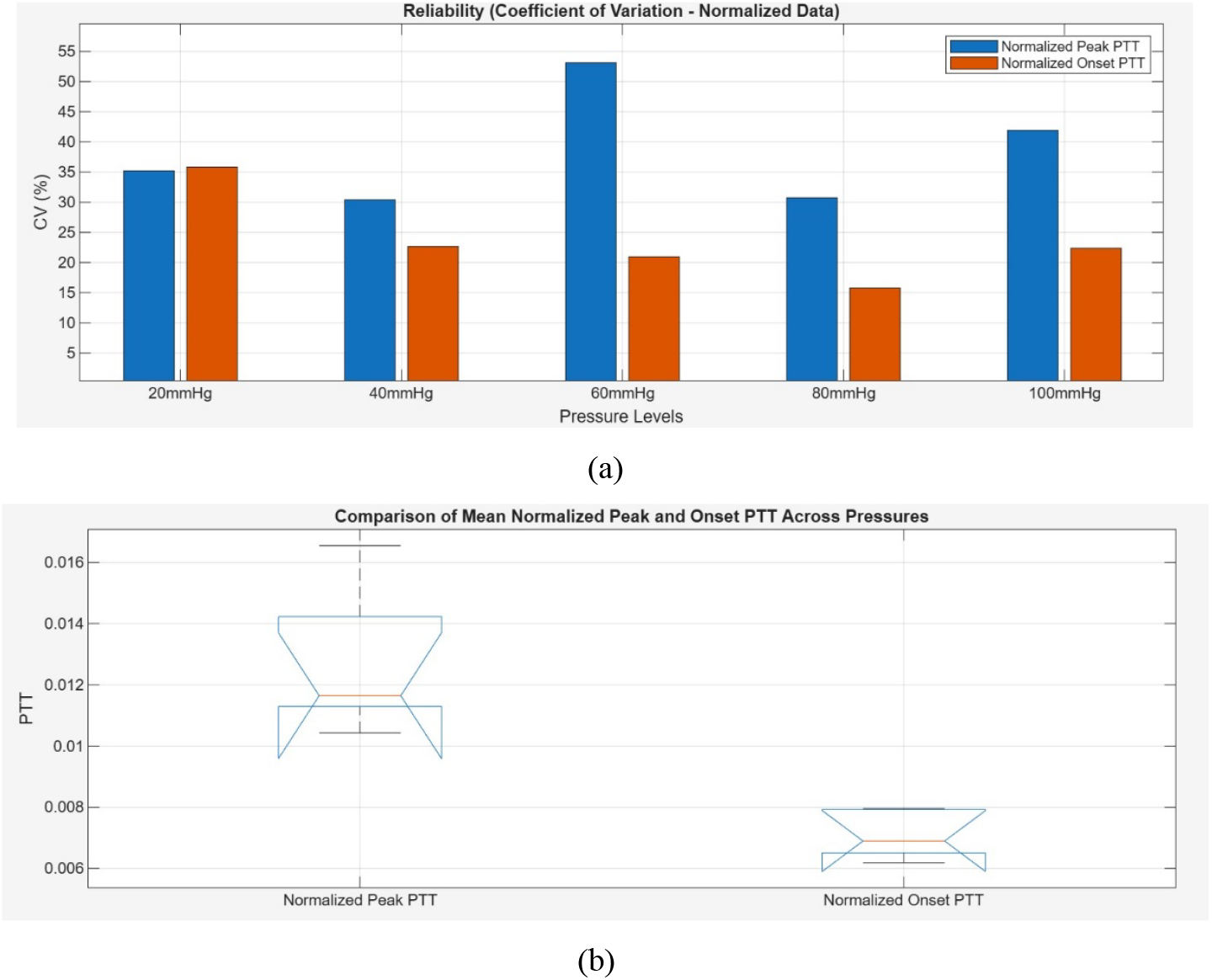
(a) Coefficient of variation (CV) of PTTpeak and PTTonset at different pressure level; (b)Normalised mean PTTonset and PTTpeak across different pressure level.

#### 3.1.2. Reliability of PTT with different pressure levels for the initial subject

A Pearson correlation coefficient is computed to check whether any relation between the time domain sequence of PTTonset and PTTpeak. This result indicates that the two measurements do not show strong linear correlation between the PTTonset and PTTpeak in this dataset. Table 4 provides the correlation index between PTTonset and PTTpeak for 20 mmHg to 100 mmHg. Since correlation measures only linear association between two sequences, it may not capture similarity in the progression of the pulse waveform over time. However, similarity between the temporal sequences of PTTpeak and PTTonset can be evaluated using the DTW method. The DTW distance is relatively stable across the pressure levels as listed in Table 5.

**Table 4:**
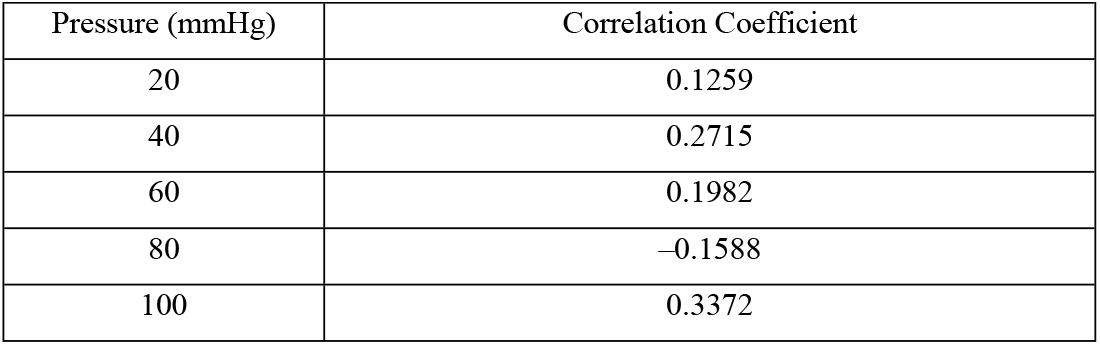
Pearson’s coefficient of correlation between PTTpeak and PTTonset.

|  |  |
| --- | --- |
| Pressure (mmHg) | Correlation Coefficient |
| 20 | 0.1259 |
| 40 | 0.2715 |
| 60 | 0.1982 |
| 80 | -0.1588 |
| 100 | 0.3372 |

**Table 5:** DTW distance between PTTpeak and PTTonset for a single participant experiment.

|  |  |
| --- | --- |
| Pressure (mmHg) | DTW Distance |
| 20 | 0.1202 |
| 40 | 0.1697 |
| 60 | 0.1943 |
| 80 | 0.2444 |
| 100 | 0.1404 |

The DTW distances ranged from 0.1202 to 0.2444 across pressure levels, showing that the time series of PTTonset and PTTpeak can be closely aligned. Although the degree of alignment varies with pressure level, the relatively small distances indicate that similar temporal patterns are present between PTTpeak and PTTonset across the tested pressures.

A comparative analysis is also performed to check the reliability between the PTTonset and PTTpeak between each pressure level. Figure 10(a) shows the confusion matrix of PTTpeak with different pressure level (20 mmHg to 100 mmHg).

**Figure 10:**
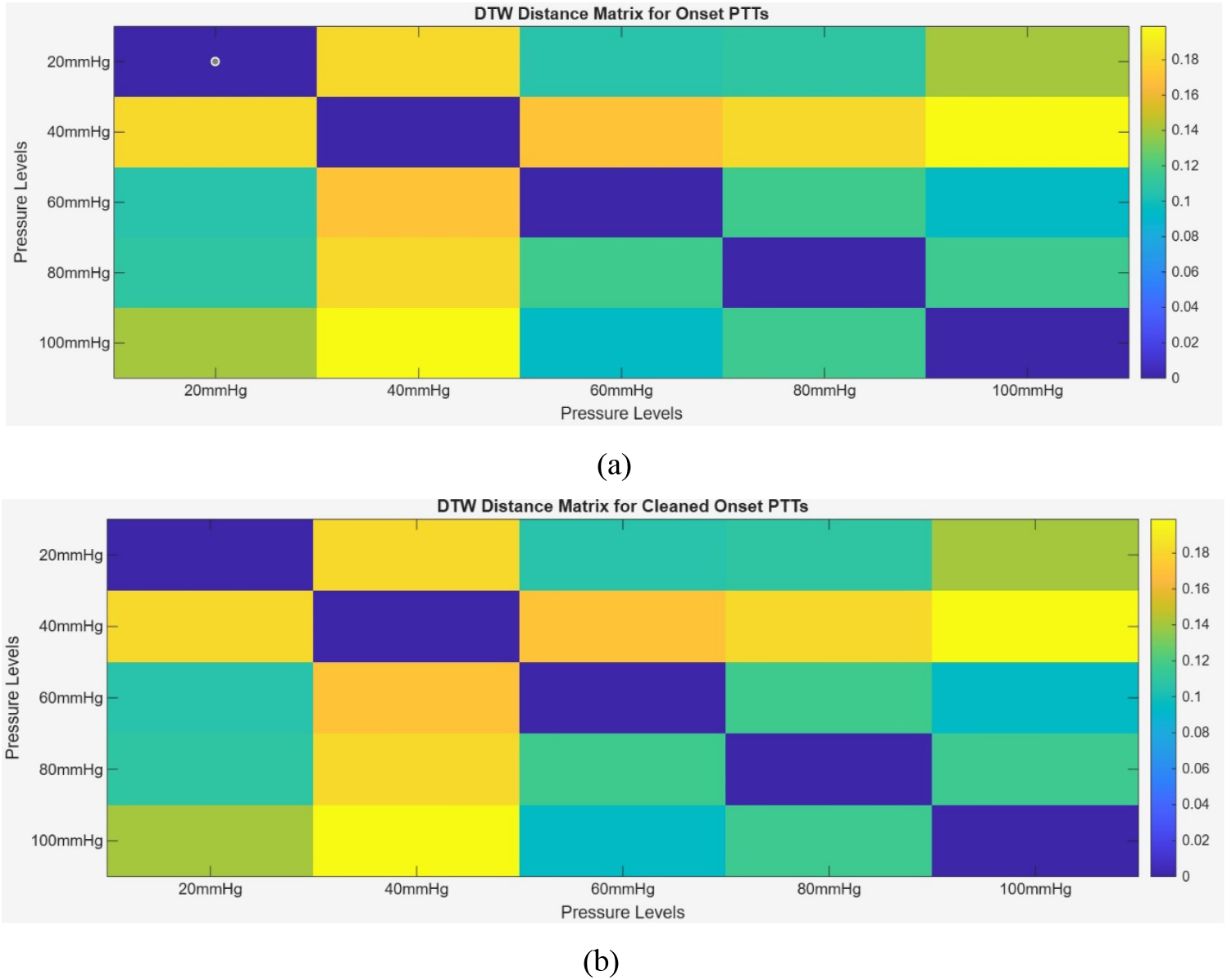
(a) Confusion matrix of DTW for PTTpeak; (b) Confusion matrix of DTW for PTTonset.

Figure 10(b) indicates the confusion matrix of PTTonset with pressure level. The experiments reveal that the time series for PTTonset and PTTpeak has higher similarity within the same pressure level. However, the similarity decreases over time when PTT compared with the neighbouring pressure level. For the single participant experiment, the measured PTT values do not appear to change strongly across the tested cuff pressure levels. However, this observation is based on a single dataset, and the effect of cuff pressure is further examined using the recordings obtained from multiple participants in the following section.

These observations from the initial subject provide a preliminary understanding of the behavior of PTTpeak and PTTonset across different pressure levels. The same analysis is extended to recordings obtained from 18 participants in the next section to validate these observations.

### 3.2. The validation Phase

#### 3.2.1. Reliability of PTT with different pressure levels among all 18 subjects

This phase evaluates whether the patterns observed in the exploratory experiment appear consistently across multiple participants and across different cuff pressure levels. To validate the above observations beyond the single subject experiment, recordings are obtained from 18 healthy subjects of age group 18 to 45 years. The data acquired is examined for the variation in PTTpeak and PTTonset. Further statistical analysis is made to observe the nature distribution PTT exhibits with 18 participants. Each dataset contains 15 sec recording at 1k Hz sampling rate consisting of 15,000 samples. PTTpeak and PTTonset values are computed from 18 subjects using the algorithm developed in MATLAB script. Table 6 represent the 18 subjects PTTpeak and PTTonset statistics on PTT count obtained using peak and onset method. Median Positive count values give a variation in the count values between onset and peak based method.

**Table 6:** Summary of PTT count Statistics for peak- and onset-based methods (N = 18).

|  |  |  |  |
| --- | --- | --- | --- |
| **Pressure Level (mmHg)** | **Method** | **Median Positive Count** | **Median Negative Count** |
| 20 | PTTpeak | 17 (16–18) | 0 (0–0) |
|  | PTTonset | 12 (10–14) | 6 (4–8) |
| 40 | PTTpeak | 18 (17–18) | 0 (0–0) |
|  | PTTonset | 15 (14–17) | 3 (2–5) |
| 60 | PTTpeak | 13 (10–15) | 0 (0–0) |
|  | PTTonset | 11 (9–12) | 2 (1–3) |
| 80 | PTTpeak | 15 (13–17) | 0 (0–0) |
|  | PTTonset | 14 (11–16) | 1 (0–2) |
| 100 | PTTpeak | 18 (17–19) | 0 (0–0) |
|  | PTTonset | 18 (17–18) | 0 (0–1) |

The PTTpeak and PTTonset of all participants with mean, SD, CV and DTW are tabulated in Table 7. The values are consolidated for different pressure levels for easy understanding of the data variation. The data obtained from the single participant reflected again across the 18 subjects.

**Table 7:** Variation of PTTpeak and PTTonset statistics among 18 participants.

| Pressure Level(mmHg) | PTTpeak Mean ± SD | PTTpeak CV (%) ± SD(%) | PTTonset Mean ± SD | PTTonsetCV (%) ± SD(%) | DTW Distance ± SD |
| --- | --- | --- | --- | --- | --- |
| 20 | 0.0102 ± 0.0008 | 46.7122 ± 10.6612 | 0.0062 ± 0.0018 | 16.8753 ± 8.1378 | 0.0459 ± 0.0301 |
| 40 | 0.0118 ± 0.0013 | 42.1450 ± 9.6112 | 0.0072 ± 0.0016 | 13.5859 ± 6.6700 | 0.0293 ± 0.0168 |
| 60 | 0.0110 ± 0.0018 | 52.4189 ± 13.2106 | 0.0071 ± 0.0018 | 8.8265 ± 5.8896 | 0.0333 ± 0.0212 |
| 80 | 0.0147 ± 0.0017 | 38.1000 ± 9.5471 | 0.0083 ± 0.0009 | 16.6381 ± 7.9107 | 0.0356 ± 0.0178 |
| 100 | 0.0116 ± 0.0009 | 39.2861 ± 9.2837 | 0.0081 ± 0.0011 | 13.1006 ± 6.5612 | 0.0382 ± 0.0197 |

As shown in Table 7, the pooled mean PTTpeak values ranged from 0.0102 to 0.0147 seconds across cuff pressures, whereas the corresponding PTTonset means ranged from 0.0062 to 0.0083 seconds. This indicates that the PTTonset is consistently lower in magnitude than PTTpeak. This difference in magnitude arises because the onset point occurs earlier in the pulse waveform than the peak, while the differences in variability and detection reliability arise from the measurement behavior observed in the experimental data.

A similar pattern is observed in the dispersion measures where maximum of 52% is reported in PTTpeak CV and 16% in PTTonset. DTW distance is also consistently lower and quantifies temporal alignment between PTTpeak and PTTonset. The mean DTW distances ranged from 0.029 to 0.046 across pressure levels, indicating similar temporal patterns between the two PTT definitions. These results indicate that PTTonset provides lower statistical variation across participants, while PTTpeak provides higher detection reliability due to fewer invalid pulse detections.

Table 8 reports the JB normality test applied to the PTTpeak and PTTonset measurements of each individual subject. For most participants the test does not reject the normal distribution assumption at 5% significance level. Only one case shows rejection of the normal distribution for the onset measurements. These results indicate that the pulse transit time distribution within individual subjects is generally close to a normal distribution.

**Table 8:** Result of Normality test for PTTonset and PTTpeak.

| Table 8: Result of Normality test for PTTonset and PTTpeak. |  |  |  |  |  |  |  |  |  |  |
| --- | --- | --- | --- | --- | --- | --- | --- | --- | --- | --- |
| Subjects | PTTpeak |  |  |  |  | PTTonset |  |  |  |  |
|  | Skew | Kurt | JB | p-value | Decision | Skew | Kurt | JB | p-value | Decision |
| Subject 1 | 0.390 | -0.99 | 3.439 | 0.179 | Normal | -0.524 | -0.49 | 2.762 | 0.251 | Normal |
| Subject 2 | 0.436 | -0.62 | 2.891 | 0.236 | Normal | -1.268 | 0.40 | 2.750 | 0.253 | Normal |
| Subject 3 | 0.299 | -1.56 | 4.409 | 0.110 | Normal | -1.658 | 3.13 | 2.294 | 0.318 | Normal |
| Subject 4 | 1.152 | 1.05 | 1.897 | 0.387 | Normal | 0.722 | -1.94 | 5.524 | 0.063 | Normal |
| Subject 5 | 1.518 | 2.61 | 1.951 | 0.377 | Normal | 0.228 | -1.81 | 4.855 | 0.088 | Normal |
| Subject 6 | 1.240 | 1.62 | 1.679 | 0.432 | Normal | -0.886 | 0.87 | 1.601 | 0.449 | Normal |
| Subject 7 | 0.834 | -1.66 | 5.101 | 0.078 | Normal | 0.115 | -2.08 | 5.392 | 0.067 | Normal |
| Subject 8 | 0.674 | 1.66 | 0.750 | 0.687 | Normal | -0.898 | 1.15 | 1.383 | 0.501 | Normal |
| Subject 9 | 1.110 | 1.12 | 1.763 | 0.414 | Normal | 0.035 | -0.84 | 3.080 | 0.214 | Normal |
| Subject 10 | 1.090 | 2.46 | 1.049 | 0.592 | Normal | -0.904 | 0.07 | 2.475 | 0.290 | Normal |
| Subject 11 | -0.515 | -0.22 | 2.385 | 0.303 | Normal | -0.990 | 2.27 | 0.926 | 0.629 | Normal |
| Subject 12 | 1.068 | -0.14 | 3.005 | 0.223 | Normal | -1.333 | 1.90 | 1.730 | 0.421 | Normal |
| Subject 13 | 1.336 | 1.40 | 2.020 | 0.364 | Normal | -0.962 | 1.51 | 1.237 | 0.539 | Normal |
| Subject 14 | -0.714 | -0.55 | 3.049 | 0.218 | Normal | -0.495 | -1.99 | 5.400 | 0.067 | Normal |
| Subject 15 | 1.337 | 2.04 | 1.682 | 0.431 | Normal | -0.586 | -0.54 | 2.893 | 0.235 | Normal |
| Subject 16 | 1.708 | 3.17 | 2.438 | 0.296 | Normal | 0.367 | -2.47 | 6.341 | 0.042 | Reject |
| Subject 17 | 1.368 | 1.64 | 1.945 | 0.378 | Normal | -0.894 | -0.17 | 2.758 | 0.252 | Normal |
| Subject 18 | 0.429 | 0.05 | 1.970 | 0.373 | Normal | -0.753 | -0.85 | 3.563 | 0.168 | Normal |

However, when measurements from all participants are combined to form pooled datasets across pressure levels, the aggregated distribution contains both within subject variation and between subject variation. The JB test applied to the pooled datasets indicates that several of these pooled measurements deviate from the normal distribution assumption. For this reason, a nonparametric Wilcoxon Signed Rank (WSR) test [34] is applied to evaluate the paired PTTonset and PTTpeak measurements across subjects. The purpose of this comparison is to evaluate whether the difference between the two measurement definitions appears consistently across subjects. This test examines whether the paired differences between PTTonset and PTTpeak measurements are systematically oriented in the same direction rather than occurring due to random variation. We note that, the purpose of using the WSR test is not to establish the ordering of waveform features, but to evaluate whether the observed difference between the two PTT definitions remains consistent across subjects and pressure conditions.

The results of the WSR test show that the paired differences between PTTonset and PTTpeak are consistently negative and statistically significant at 20, 40, 80, and 100 mmHg at 95% confidence level. At 60 mmHg the difference is not statistically significant at 95% confidence level (but at 90% confidence level). These results state that the difference between the two PTT definitions appears consistently across most subjects and pressure levels.

In order to describe the magnitude of the paired differences between PTTonset and PTTpeak, the median difference (PTTonset minus PTTpeak) is also computed at each pressure level. The median differences are negative for all pressures, indicating that PTTonset values were consistently lower than PTTpeak values. In addition, the majority of participants at every pressure level show that PTTonset values are lower than PTTpeak values. This behavior confirms that the observed differences appear consistently across subjects and are not driven by isolated subjects.

The onset PTT showed lower variability across all participants and cuff pressures, which supports its stability, as shown in Table 7. However, the PTTpeak demonstrated a lower error rate, particularly at lower cuff pressures where onset detection was occasionally affected by motion or waveform distortion. This observation suggests that reliability depends on both measurement stability and detection robustness. The onset method provides lower variability, while the peak method is less sensitive to waveform distortion. The low DTW distance values observed across participants and pressure levels indicate that the PTTonset and PPTpeak share highly similar temporal patterns even when their absolute values differ. This finding suggests that both definitions capture the same underlying behavior but represent it at different time reference points within the waveform.

In addition, a Friedman test was conducted to examine whether pulse transit time varied across the five cuff pressures. The analysis showed a statistically significant pressure effect for both PTTpeak and PTTonset. The chi-squared statistic with 4 degrees of freedom was 42.77 with p-value less than 0.001 and Kendall’s coefficient of concordance (Kendall’s W) 0.594 for PTTpeak, while the corresponding values were 14.89, p-value 0.0049, and Kendall’s W 0.207 for PTTonset. Post-hoc WSR tests with Bonferroni correction showed that several pressure pairs differed significantly for PTTpeak, particularly those involving 80 mmHg. For PTTonset, significant differences were observed only between 20 mmHg and the higher pressure levels (80 and 100 mmHg). These results indicate that PTT values vary across cuff pressures, with a stronger effect observed for peak measurements.

From a practical perspective, models designed to track relative changes in blood pressure over time could use either PTT definition with minimal loss of temporal fidelity. However, for estimating absolute blood pressure values, calibration to account for the systematic offset between onset and peak timings may be necessary. The present study evaluates the measurement behavior of the two PTT definitions rather than constructing a direct blood pressure prediction model.

## 4. Conclusion

The PTT values are measured using two pressure sensors placed along the radial artery to examine the statistical behavior of pulse transit time computed using peak and onset definitions. The PTTonset shows 8.8% as lowest coefficient of variation as compared to PTTpeak. The standard deviation is also minimum with PTTonset with a value 0.006. In terms of mean and standard deviation, PTTonset has better stability in the reading as compared to PTTpeak. However, during the computing complexity, PTTonset needs more computational resource and time as compared to PTTpeak. PTTpeak can be easily computed and with better positive pulse count. PTTonset may have negative pulse count as drawback due to motion artifact and is reflected in the computation of wrong time index point on the pulse. However, PTTonset values show lower variability than PTTpeak. The correlation coefficients are small in magnitude, indicating that the two PTT definitions do not exhibit strong linear association in their time sequences across the recorded pressure levels. However, time series of PTTonset and PTTpeak show comparable temporal patterns using DTW. A similarity is found between PTTonset and PTTpeak with DTW distances ranging from 0.029 to 0.046 across tested cuff pressures, with the lowest value at 40 mmHg indicating that the two sequences can be matched each other with very little time warping. The JB normality test was applied to PTTpeak and PTTonset values for each subject to assess the underlying data distribution. The results indicate that PTTpeak values are normally distributed for almost all subjects, as evidenced by p-values greater than 0.05.

## Data Availability

The experimental protocol involving human participants was reviewed and approved by the Institutional Ethics Committee of NMAM Institute of Technology (NMAMIT), Nitte (Deemed to be University), Karnataka, India (Approval Year: 2020). The physiological data used in this study were collected in 2022 under the approved protocol. All participants provided informed consent prior to data collection, and all procedures were conducted in accordance with the principles of the Declaration of Helsinki.

## Declaration

### Funding

The part of entitled research work was funded by the Indian Council of Medical Research. Authors also reports that the article processing charges for this publication were supported by NITTE (Deemed to be University).

### Data availability

The datasets generated and analysed during the current study are provided as supplementary material associated with this article

### Author Contributions

S.R.M. conceived the idea, designed the experimental setup, conducted the experiments and data collection, prepared the initial draft of the manuscript, and secured funding for the study. D.K. performed the statistical analysis and critically reviewed and revised the manuscript. Both authors reviewed and approved the final manuscript

## Notes

### Competing Interest Statement

The authors have declared no competing interest.

### Clinical Trial

NA

### Funding Statement

Yes

